# Quantification and optimization of travel time for ethnic minority populations in cancer clinical trials

**DOI:** 10.1101/2024.05.29.24308033

**Authors:** H Lee, JG Trevino, MB Terry, K Winkfield, T Janowitz

## Abstract

Of all minority racial and ethnic groups, Hispanic populations are most under-represented in trials compared to the general US population. Transportation and socioeconomic burdens are two important inter-related quantifiable and modifiable variables associated with decreased clinical trial participation for underrepresented populations. In this study, Hispanic population sizes and socioeconomic deprivation indices of catchment areas within simulated 30-minute driving distances from all major U.S. cancer trial sites (N=78) and all U.S. hospitals (N=7,623) were calculated using OpenStreetMap and U.S. census data. In the proximity of major trial sites Hispanic ethnicity representation varied across a wide range (64% to 2%) and Hispanic populations were underrepresented compared to the national average in almost 2/3 of the sites (n=50). The cities with the highest number of hospitals identified with catchment populations of >60%, >40% or >20% Hispanic representation were San Antonio TX, Houston TX, and New York NY respectively. Data-driven analyses can quantify and optimize measurable factors associated with decreased clinical trial participation for under-represented populations and may aid selection of trial sites to enable participation.

## INTRODUCTION

Clinical trials should reflect the true diversity of the population^1^. Of all minority racial and ethnic groups, Hispanic populations are most under-represented in trials compared to the general US population^2^. In landmark trials leading to US Food and Drug Administration (FDA) approval of oncology drugs, Hispanic ethnicity is underrepresented by less than half of expected numbers, despite the growth of Hispanic population sizes and their increased relative cancer incidence and mortality burden^3^. Transportation and socioeconomic burdens are two important inter-related quantifiable and modifiable variables associated with decreased clinical trial participation for underrepresented populations^4^. Travel burdens for such populations can be quantified and potentially reduced, as demonstrated for patients who identify as Black/African American in the U.S. census^5^. Here we conduct an analysis to quantify and optimize trial access by travel distance for Hispanic populations across the U.S.

## METHODS

Major U.S. cancer trial sites (N=78) were collated using the National Cancer Institute Comprehensive Cancer Center list and filtering for U.S. based sites from the nature-index top 100 cancer research healthcare institutions (Supplement1). These 78 major U.S. cancer trial sites were listed in 94% of all U.S cancer trials between 2012-2022. Hispanic population sizes and socioeconomic deprivation indices of catchment areas within simulated 30-minute driving distances from all major U.S. cancer trial sites (N=78) and all U.S. hospitals (N=7,623) were calculated using OpenStreetMap and U.S. census data (Supplement1). The 30-minute time cut-off was based on published travel distance durations that maintained higher patient enrollment rates (Supplement1). Sensitivity analyses to identify potential satellite hospitals with reduced travel burden for Hispanic populations were conducted by filtering all hospitals for catchment populations with over 20-, 40- or 60% Hispanic representation and with a catchment population size large enough to recruit at least 500 cancer patients using the national average cancer prevalence and trial enrollment rate (Supplement1). All analyses and map visualizations were conducted in R studio (v2022.02.1+461) and R (v4.1.3).

## RESULTS

Major U.S. cancer trial sites (N=78) showed a wide range of Hispanic ethnicity representation (64% to 2%) within a 30-min single direction commute (Figure 1A), with Hispanic populations underrepresented compared to the national average in almost 2/3 of the sites (n=50). Overlay of longitudinal data showed increasing Hispanic representation in the catchment areas of most U.S. cancer research sites over the last 15 years (70/78 sites, Figure 1A). Sensitivity analyses were conducted to identify existing hospitals with catchment populations that represented the overall national proportion of Hispanic populations (>20%) or more (>40%, >60%), as well as having a catchment population size large enough to successfully enroll 500 cancer patients (Figure1B; Supplement1). The cities with the highest number of hospitals identified with catchment populations of >60%, >40% or >20% Hispanic representation were San Antonio TX, Houston TX, and New York NY respectively. Mapping of identified hospitals within their surrounding city areas cartographically highlights their spatial proximity to urban tracts with high ethnic diversity (Figure 2 A, C, D) and socioeconomic diversity (Figure 2B) whilst also being within 30-min commuting borders from pre-existing major cancer trial sites^A^.

**Figure 1.**
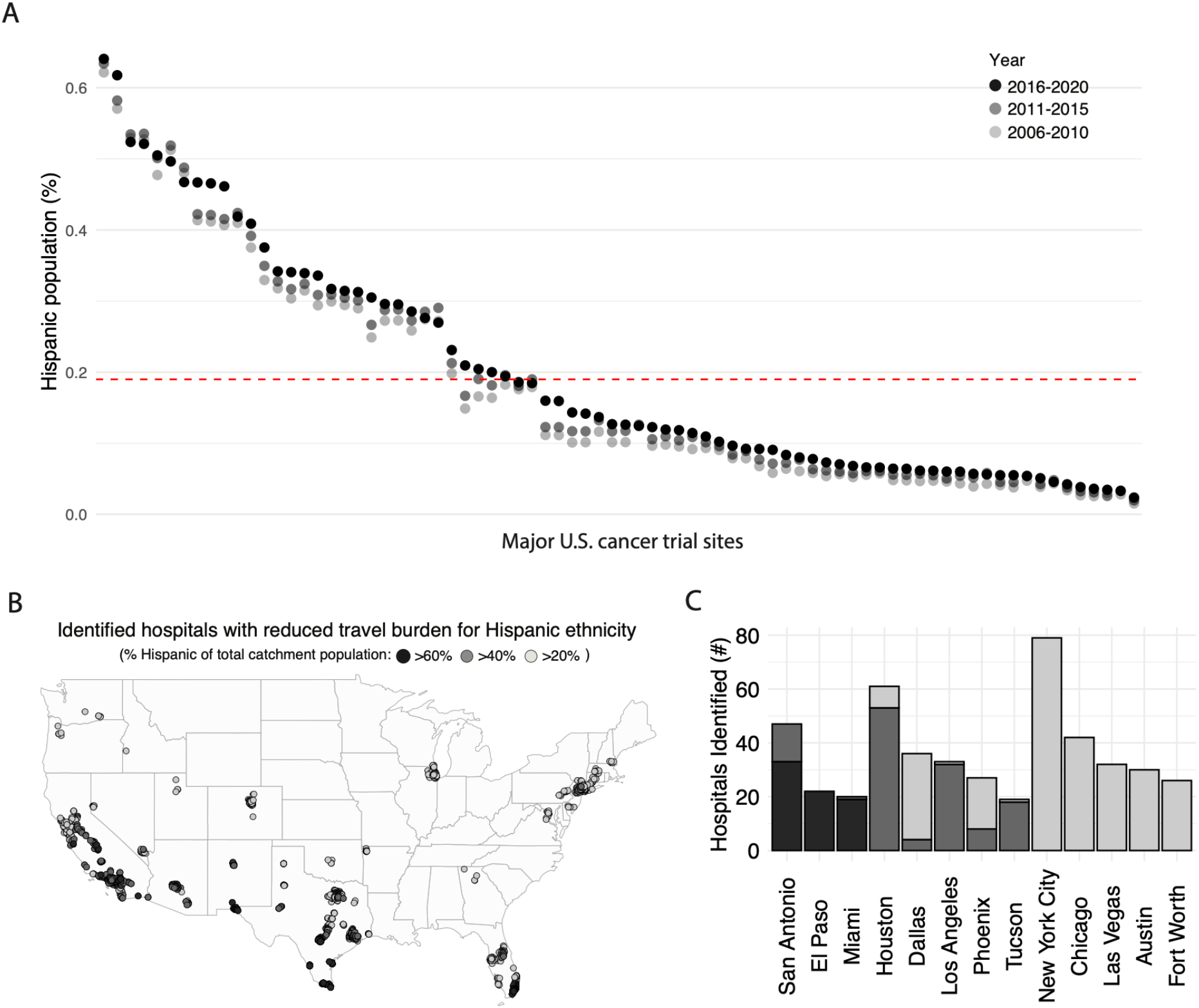
Quantification and sensitivity analyses of Hispanic representation for commuting-populations of high-volume U.S. cancer trial centers and all U.S. hospitals. A, Dot plot showing the representation of Hispanic populations living within 30-minute commuting distance from each high-volume cancer trial site in the U.S. (% of total commuting population). Trial sites are ordered on the x-axis by descending order of % representation of Hispanic populations for their respective commuting populations. Quinquennia are represented as follows: 2016-2020, 100% opacity; 2011-2015, 50% opacity; 2006-2010, 25% opacity. The red dotted line depicts the current population proportion of self-identified Hispanic individuals in the U.S. (19%). B, Map of U.S. based hospitals identified as having reduced travel burden for Hispanic ethnicity populations as defined by % representation within a 30-minute commuting distance catchment area. (Hospitals with >20%, >40% or >60% Hispanic representation colored as light-grey, grey, dark-grey respectively). C, Stacked bar graph showing the top 13 cities with the highest number of hospitals identified as having a catchment population with an above average Hispanic representation of >20%, >40%, or >60% (light-grey, grey, dark-grey respectively).

**Figure 2.**
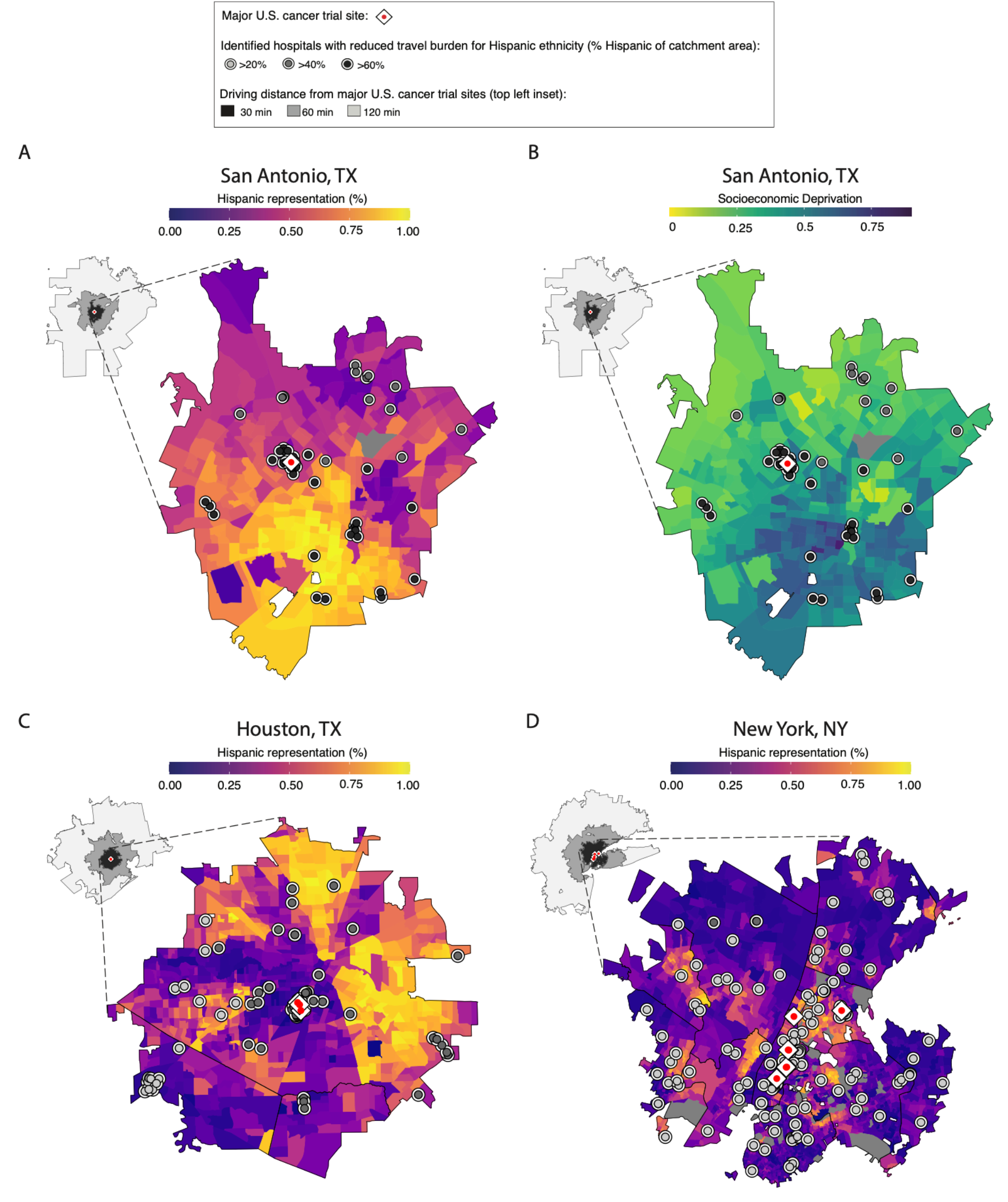
Maps showing Hispanic representation and socioeconomic deprivation with overlay of high Hispanic representation hospitals and commute boundaries from existing high volume cancer trial sites. A, C and D, Map of commuting-populations within 30-minute travel time to existing cancer research hospitals (white vertical rhombus with superimposed internal red dot) in cities San Antonio TX, Houston TX, New York NY respectively. These cities were identified as containing the highest numbers of hospitals with high Hispanic representation within their catchment areas in Figure 1C. Top left inset (increasing shades of grey) outline the areas that fall under 120-, 60-, or 30-minute one-way commute times, respectively, from the high-volume cancer trial sites within those cities. Tracts are colored by Hispanic representation (with lighter-yellow tracts showing greater proportion of residing Hispanic populations). Existing hospitals identified through sensitivity analyses in Figure 1 for high Hispanic representation within their 30-minute catchment areas are shown by circles with black-white-black triple borders with increasingly dark internal coloring to represent >20%, >40%, >60% Hispanic representation. B, Map of commuting-populations within 30-minute travel time to existing cancer research hospitals (white vertical rhombus with superimposed internal red dot) in San Antonio TX. Analogous maps for New York NY and Houston TX are previously published ^5^. Top left inset (increasing shades of grey) outline the areas that fall under 120-, 60-, or 30-minute commute times, respectively, from the cancer research hospitals within the city. Tracts are colored by census level socioeconomic deprivation indices. Darker-colored tracts with deprivation index values closer to 1 represent greater socioeconomic deprivation. Overlays of existing high-volume cancer research sites and potential satellite hospitals with high Hispanic representation in their catchment areas are provided as for other panels.

## DISCUSSION

Data-driven analyses can quantify and optimize measurable factors associated with decreased clinical trial participation for under-represented populations^5^. This study shows that the majority of the major U.S. cancer trial sites (N=78; listed in 94% of all U.S. cancer trials between 2012-2022) underrepresent Hispanic populations within their catchment areas (Figure1a), but that some sites serve areas with high Hispanic population representation, and highlights potential satellite hospital site locations that could be utilized to reduce travel burdens for Hispanic patient populations in proximity to pre-existing major cancer research hospitals (Figure2).

We acknowledge that the U.S. census categorization of all ethnicities into Hispanic and Non-Hispanic does not provide high granularity for the very heterogenous group of individuals of varying ethnicities and ancestries living within the U.S. Also, other non-travel related factors such as language and cultural awareness can also impact the enrolment and retention of patients and must be considered and addressed to improve equitable inclusion of Hispanic patients into clinical trials^6^.

## Data Availability

All data produced in the present study are available upon reasonable request to the authors.

## ACKNOWLEDGEMENTS

We acknowledge funding from Cold Spring Harbor Laboratory and Northwell Health (HL and TJ).

## Footnotes

A Extended maps for socioeconomic diversity in other cities are available in prior manuscript.^5^

