## Extended Methods for "Quantification and optimization of travel time for ethnic minority populations in cancer clinical trials"

### Analysis and optimization of equitable U.S. cancer clinical trial center access by travel time

#### eMethods

##### Data sources

A list of major U.S. cancer clinical trial centers (N = 78) were collated from two data sources, the nature-index top 100 cancer research healthcare institutions list[1] and non-laboratory based National Cancer Institute Comprehensive Cancer Centers[2]. Trial volume for each institution between 2012-2022 was queried on the national trial registry (nct.gov) filtered by age (>18), location (U.S.), trial type (phase 1, 2, 3) and recruitment status (excluded if 'terminated' or 'withdrawn'). Trials with more hospital sites registered than number of patients were excluded. The major U.S. cancer trial centers (N = 78) were found to be listed in 94% of all U.S. cancer trials.

The geographic locations of all hospitals in the U.S. (N=7,623) were taken from The Homeland Infrastructure Foundation-Level Data.[3]

Ethnicity representation data were obtained by retrieving self-identified ethnicity population counts from the U.S. census American Community Survey between 2006-2010, 2011-2015 and 2016-2020 using the *tidtycensus* R package[4]. Ethnicity category variables collected through the census were either Hispanic or Latino (B03001_003) or Non Hispanic or Latino (B03001_002). The U.S. census bureau provided geographical crosswalk data file (https://www2.census.gov/geo/docs/maps-data/data/rel2020/tract/tab20_tract20_tract 10_natl.txt) was downloaded to correlate historical 2010 census tracts to updated 2020 census tracts. This resulted in re-identification of 96.5% of called 2020 census tracts within historical census data.

Deprivation indices[5] were calculated from U.S. 2020 census American Community Survey data collected using the *tidycensus* R package[4] using variables for fraction vacant housing (B25002_003), assisted income (B19058_002), health insurance status (B27010_{017,033,050,066}), median income (B19013_001), high school education (B15003_017-B15003_025), and poverty level (B17001_002) per census tract.

30-minute catchment population simulation

The *osrm* R package (Open StreetMap[6,7]) was first used to simulate driving times to the centroids of all U.S. census tracts located within a 200km flying-distance to chosen hospital or cancer research sites. Catchment populations were defined as the combination of all U.S. census tracts with centroids located within 30-minute one-way driving distance away from chosen hospital or cancer research site locations. The time cut-off was based on published travel distance duration cut-offs that maintained higher patient enrollment rates[8].

Sensitivity Analyses

First, the proportional representation of Hispanic ethnicity within 30-minute driving distance to every hospital in the U.S. was calculated by dividing the sum of Hispanic populations living in census tracts within the simulated 30-minute driving times for each hospital site by the sum of its total 30-minute catchment population size. Thereafter, all hospitals were filtered for Hispanic representation of at least the national average (>20%) or more (>40%, >60%).

Second, all remaining hospitals with total catchment populations below a threshold catchment population size were excluded. The threshold catchment population size was calculated to be of approximately sufficient size to recruit 500 cancer patients into a clinical trial. This cut off was in the upper range of the mean number of patients recruited for interventional phase 1, 2, or 3 trials between 2012–2022 from the 78 most active cancer clinical trial sites as described in the data sources section (mean 126, 250, 1,321; standard error 3, 13, 48 respectively). The threshold population size was calculated by dividing the desired number of enrolled patients by the prevalence of all cancers in the U.S. population (5.2% in 2020)[9], and by the national average enrollment rate of adult cancer patients into clinical trials (6.3% in 2021)[10]. Calculated in this manner, the threshold size represents a reasonable expectation of the minimum catchment population size required to recruit sufficient numbers of participants, given current cancer incidence and enrollment rates for an active trial site.

Software and Packages

All analyses were conducted and visualized in R studio (v2022.02.1+461) and R (v4.1.3) using *Tidyverse*[11] and *ggplot2.* Two-group estimation plots were producsed using *dabestr*[12]. Topologically Integrated Geographic Encoding and Referencing system (TIGER)/Line shapefiles of the legal boundaries of U.S. census tracts and counties were collected using the *Tigris* R package[13].


1. Top 100 healthcare institutions in cancer research | Nature Index 2020 Cancer | Supplements | Nature Index. Accessed October 2, 2022. https://www.nature.com/nature-index/supplements/nature-index-2020-cancer/tables/healthcare
2. NCI-Designated Cancer Centers - NCI. Accessed October 2, 2022. https://www.cancer.gov/research/infrastructure/cancer-centers
3. HIFLD Open Data. Accessed October 2, 2022. https://hifld-geoplatform.opendata.arcgis.com/search?collection=Dataset
4. Walker K, Herman M. tidycensus: Load US Census Boundary and Attribute Data as “tidyverse” and ’sf’-Ready Data Frames. *R package version 123*. Published online 2022. Accessed October 3, 2022. https://walker-data.com/tidycensus/ )
5. Brokamp C, Beck AF, Goyal NK, Ryan P, Greenberg JM, Hall ES. Material community deprivation and hospital utilization during the first year of life: an urban population-based cohort study. *Ann Epidemiol*. 2019;30:37-43. doi:10.1016/J.ANNEPIDEM.2018.11.008
6. OpenStreetMap. Accessed October 2, 2022. https://www.openstreetmap.org/about
7. Giraud T. osrm: Interface Between R and the OpenStreetMap-Based Routing Service OSRM. *J Open Source Softw*. 2022;7(78):4574. doi:10.21105/JOSS.04574
8. Legge F, Eaton D, Molife R, et al. Participation of patients with gynecological cancer in phase I clinical trials: Two years experience in a major cancer center. *Gynecol Oncol*. 2007;104(3):551-556. doi:10.1016/J.YGYNO.2006.09.020
9. Cancer of Any Site — Cancer Stat Facts. National Cancer Institute, Surveillance, Epidemiology, and End Results Program. Accessed October 23, 2023. https://seer.cancer.gov/statfacts/html/all.html
10. Unger JM, Fleury M. Nationally representative estimates of the participation of cancer patients in clinical research studies according to the commission on cancer. *https://doi.org/101200/JCO20203928_suppl74*.
11. Wickham H, Averick M, Bryan J, et al. Welcome to the Tidyverse. *J Open Source Softw*. 2019;4(43):1686. doi:10.21105/JOSS.01686

12. Ho J, Tumkaya T, Aryal S, Choi H, Claridge-Chang A. Moving beyond P values: data analysis with estimation graphics. *Nature Methods 2019 16:7*. 2019;16(7):565-566. doi:10.1038/s41592-019-0470-3
13. Walker K. Tigris: An r package to access and work with geographic data from the us census bureau. *R Journal*. 2016;8(2):231-242. doi:10.32614/RJ-2016-043
